# Patient-reported outcomes relevant to post-discharge trauma patients in urban India

**DOI:** 10.1101/2024.02.20.24302971

**Authors:** Siddarth David, Martin Gerdin Wärnberg, Trauma life support training Effectiveness Research Network (TERN) collaborators

## Abstract

Trauma is a major global health burden with long-term consequences to patients. Patient-reported outcomes (PROs) are increasingly being recognized and utilised to improve trauma care. It is imperative to identify the PROs that are most relevant, applicable, and suitable to a context. However, there is limited research on PROs in trauma care in low- and middle-income countries, like India, that bear a disproportionate burden of global trauma. This paper aims to examine PROs considered to be most relevant for trauma patients after discharge in the context of urban India.

We conducted semi-structured qualitative interviews of 11 adult post discharge trauma patients, across demographic and injury groups, and two persons working with trauma patients from five tertiary-care public hospitals in the Indian cities of Mumbai and Kolkata. We performed thematic analysis to identify themes within the participants responses on PROs important to them based on their experiences.

Four themes emerged in the analysis of the participant interviews. The need for full physical functioning, the need to address psychological consequences, the need to alleviate economic cost of trauma, and the need for social interactions. Outcomes related to these themes were most relevant to the participants.

The findings of this paper can help researchers and clinicians select appropriate PROs, based on the four themes, for trauma research and to improve trauma care in urban India. Future research should focus on PROs relevant to specific demographic and injury groups in India and other LMICs.

## Background

Trauma results in around four million deaths across the globe and is the top cause of death among adolescents and young adults (1). Additionally, nearly one billion people worldwide, survive trauma and live with consequences of trauma, especially in the 20 to 69 age group, requiring post-discharge continuum of care through the healthcare system (2). Low- and middle-income countries (LMICs) bear a disproportionate share of this trauma burden (3). Physiological functioning, socioeconomic outcomes, psychological well-being, and overall quality of life remain affected post-discharge and improving them to pre-injury levels are part of the recovery process for trauma patients (4–8). The healthcare system can play a crucial role in this recovery process and ameliorate these consequences of trauma (9). This makes it imperative that the provision of care should be based on patient requirements.

Integrating patient perspectives to better meet patient needs has been widely accepted in healthcare practice and policy (10,11). Patient-reported outcomes (PROs) in healthcare care, including trauma care, can provide clinicians and researchers with information to incorporate patient perspectives, improve patient care, and inform clinical practice and consequently health policy (12–14). Though there has been an increase and wide use of PROs in trauma care research over time, such research was the lowest in trauma compared to other disease groups (15,16). Additionally, the PROs measured are not always relevant or important to the needs of trauma patients (17).

A key characteristic of using PROs is to identify and measure outcomes that are relevant and of consequence to patients and their caregivers (18). PROs relevant to patients can differ across countries and socioeconomic groups (22,23). Using PRO measurement tools based on the needs and outcomes most important to patients in a particular setting can improve the acceptability of the tool among patients, capture outcomes relevant to them, and help the healthcare system improve to better meet their expectations (19–21). Consequently, using relevant PROs will guide clinicians and researchers in selecting standardised tools from the available PRO measurement tools that are appropriate to trauma patients in their settings.

There are many tools to measure PROs, but existing evidence on using these PROs to measure trauma care outcomes comes largely from high income countries (HICs) (24,25). LMICs bear a greater trauma burden, yet have inadequate research on PROs in trauma care; and it is crucial to identify the outcomes most relevant, applicable, and suitable to a context. Accordingly, this paper aims to examine PROs considered to be most relevant for trauma patients after discharge in the context of urban India.

## Methods

### Design

To examine PROs relevant to trauma patients in urban India, we performed thematic analysis of semi-structured interviews of discharged trauma patients and persons working with such patients. Using such a qualitative approach can extract participant responses on PROs that are important to them (26,27). Semi-structured interviews were selected to elicit descriptive accounts from the participants with the opportunity to probe particular outcomes brought up (28).

### Setting

India accounts for more than one-fifth of the global trauma burden and trauma contributes for one-tenth of the DALYs (29). The interviews were conducted in two major cities in India: Mumbai and Kolkata. These cities are large urban agglomerations with a population of 18.4 million and 14.1 million, respectively and together report around 9,000 trauma-related deaths each year (30). Both cities have a network of private and public healthcare facilities, with low- and middle-income groups using the public healthcare facilities, mainly for tertiary care (31,32). The sites of participant recruitment were five large public tertiary-care teaching hospitals in these two cities.

### Participants

This study was part of the community consultation initiatives of a pilot multicenter cluster randomised trial to study the feasibility of comparing the effect of trauma life support training programs on patient outcomes (ClinicalTrials.gov NCT05417243) (33). We included all patients above the age of 15 admitted with a history of trauma in the five hospitals. These patients included different mechanisms of trauma such as transport injuries, falls, and intentional injuries and with various anatomical injury distributions. We included both males and females in different age groups to improve the representativeness of the. The participants were those living in either Mumbai or Kolkata. We contacted the patients after three months post their discharge so that it would give them sufficient time to identify PROs most relevant to them based on their lived experience. We also interviewed people engaged with trauma patients such as hospital social workers, who help trauma patients in their recovery process.

### Data collection instruments

We developed the interview guide to obtain responses from the participants on PROs, which they considered were important for trauma patients in their recovery after discharge. It was developed through discussion with the research team that was composed of clinicians and researchers working with trauma patients in urban India. To capture relevant PROs, we included questions on the effect of trauma on their lives or lives of trauma patients; the functional, social, and economic challenges faced after discharge; and the challenges that they or trauma patients would prioritise the most. After checking for quality, we piloted the guide among the trauma patients to assess for acceptability and appropriateness for eliciting adequate responses and then revised it accordingly. Throughout the data collection process, the guide was regularly reviewed, to make modifications based on the nature of responses by the participants. The interview guide is available in the Supplementary Materials.

### Data collection

As patients were enrolled for the pilot cluster randomised trial research, their consent was sought to be interviewed post discharge by explaining in detail the purpose of the study. They were not offered any incentives or pressured to be part of the study. They were assured of confidentiality and anonymity of their responses and participation. We contacted the selected patients by telephone and after expressing willingness to be interviewed we scheduled a convenient time for the interview. The potential persons working with trauma patients such as hospital-based social workers, were contacted explaining the aim of the study and if they agreed an interview was scheduled as per their convenience.

Before the interviews, the purpose of the interviews and the confidential and anonymous nature of the interviews was reiterated. It was also clarified that they did not have to answer any questions they chose not to, they could end the interview at any point, and they could take back their consent to interview at any point. The interviews were conducted in the local languages spoken in Mumbai and Kolkata—Hindi, Marathi, Bengali, and English—based on the participants’ preference. After obtaining the consent of the participants the interviews were audio recorded. In case a participant refused to be audio recorded, detailed notes were taken during and after interviews to capture the responses of the participant. A few patients were interviewed via telephone due to inability to conduct face-to-face interviews because of the lack of a convenient time or place given the work and personal commitments of the participants.

An interview typically lasted between 30-40 minutes and was usually conducted at the participants’ home or office. All efforts were made to ensure the participant was alone and there were minimal disturbances during the interview. In case of an incapacitated patient, the primary caregiver would be asked to respond or fill in details on behalf of the patient. Apart from the recordings and written notes on the responses, field notes were maintained to document observations or descriptions relevant to the interview. The team members collecting the interviews were trained on how to conduct the interviews while adhering to robust scientific and strict ethical principles. Any unanticipated challenges during the interviews were immediately discussed with the research team and addressed. We decided the final number of interviews, by consensus within the research team that data saturation had been reached (34).

### Data management

We translated and transcribed the interviews verbatim and field notes to English. We removed all identifiable personal information from the transcripts and field notes, maintaining and storing only anonymized versions to be used for analysis. They were stored securely by SD, with limited access to only those research team members involved in analysis.

### Data analysis

The anonymized transcripts and notes were analysed using thematic analysis to identify emerging themes within the PROs deemed most relevant by trauma patients post discharge. Using a structured approach, the transcripts and notes were examined to develop codes that identified and summarised key concepts and then grouped into categories and themes (35,36). The process of identifying and coding was carried out deductively, based on a previous literature that was used to develop the interview guide as well as inductively, based on themes that came through the transcripts and notes. SD derived the initial codes and themes, which were then reviewed and discussed by the research team. A table with codes, categories, and themes is available in the Supplementary Materials.

### Techniques to enhance trustworthiness

We maintained an audit trail documenting all the decisions taken and modifications made during the study period. The data collectors also maintained a reflexive journal of the process. We periodically discussed these documents along with the transcripts and notes to improve the trustworthiness of this study (36).

### Researcher characteristics and reflexivity

The data collectors who conducted the interviews have been working on different research projects with trauma patients for over five years. They have intimately interacted with trauma patients in Mumbai and Kolkata right from the point of admission, through their hospital stay and discharge, and post discharge follow-up. SD, who supervised the data collectors, has worked in public health research—using qualitative, qualitative, and mixed methods—for over a decade, with his doctoral work focusing on the lived experience of trauma patients in urban India. The other members of the research team have been working with trauma patients and engaged in trauma-based research projects for up to 30 years in India and other global settings.

### Ethics

We obtained ethical approval for this study, as part of the larger pilot study, at all five health facilities (Approval No: HIEC/003/FEB-22/0/RP/03/02-22, Dated: 11 Feb 2022; IEC(II)/OUT/134/2022, Dated: 8 Feb 2022; Ref No.MC/KOL/IEC/NON-SPON/1217/11/21, Dated: 31 Nov 2021; NRSMC/IEC/93/2021, Dated: 29 Dec 2021; IEC/214/22 (Ref: IEC/40/22), Dated: 20 May 2022)). We sought and recorded informed consent from all the participants before the interview and permission was taken for audio recording. When contacting a potential participant, all efforts were taken to ensure that the interviews were held at their convenience, at a place and time comfortable to them to be as unobtrusive to the daily schedule of the participants. Almost all participants preferred having the interviews at their homes or workplaces, giving us an opportunity to understand the context of the participants while helping them be comfortable in a familiar setting. However, this meant potential distractions and interruptions by family members and colleagues, which we tried to minimise. Yet, such interruptions were unavoidable, which may have affected the responses of the participants.

We ensured that participants were aware that their participation was completely voluntary and they were free to refuse and withdraw at any point of the interview. With patients we assured them that their non-participation or withdrawal would not affect their current or future treatment at the health facility they were admitted in. With non-patient participants, who were primarily employed in the hospital they worked in, we assured them that their views or opinions would not be disclosed or shared with any other hospital staff. With all participants, we clarified that while they will not be directly benefited by participating in interviews, the potential findings from this study may help improve healthcare for trauma patients in the future.

Along with training the data collectors to be ethical and sensitive in their conduct of the interviews, we held debriefing sessions after the interviews as a forum for them to discuss any ethical concerns or challenges they encountered during the interviews and ways to address them. Anticipating the possible psychosocial toll on participants from recounting the challenges faced by them due to the trauma, a list of relevant organisations and public hospital departments were kept by the data collectors to share with the participants for any support they may need. The data collectors were asked to keep a list of organisations and departments in the participating hospitals that could help the participants with some of the challenges raised during the interviews. Utilising somebody’s time and experiences without them being directly befitted is an ethical concern. However, we feel that the findings of this study may help inform policy and practice to improve trauma care services in the future that can potentially benefit everyone in the long run.

## Results

### Participant Characteristics

A total of 13 participants were interviewed for this paper. 11 were trauma patients and 2 were persons working with trauma patients during their recovery process—medical social workers at the hospital. 10 participants were interviewed face-to-face while three of the participants preferred having the interview over telephone. A brief description of the demographic, injury profile, and job description of the participants is shown in Table 1 and Table 2.

**Table 1:** Participant Profile.

| ***Patients*** |  |
| --- | --- |
| ***Injury description*** | ***Length of hospital stay*** |
| **Railway injury with lower limb amputation** | 3 months |
| **Fall with polytrauma** | 1 month |
| **Road traffic injury with polytrauma** | 1.5 months |
| **Assault with abdominal injury** | 1 |
| **Fall with polytrauma** | 1.5 months |
| **Road traffic injury with traumatic brain injury (TBI)** | 0.5 months |
| **Fall with polytrauma** | 1 month |
| **Road traffic injury with polytrauma and spinal cord injury (SCI)** | 2 months |
| **Fall with polytrauma** | 0.5 months |
| **Road traffic injury with polytrauma** | 0.5 months |
| **Road traffic injury with polytrauma** | 0.25 months |
| ***Working with trauma patients*** |  |
| ***Role*** | ***Years working*** |
| Medical Social Worker in the hospital | 14 |
| Medical Social Worker in the hospital | 23 |

**Table 2:**
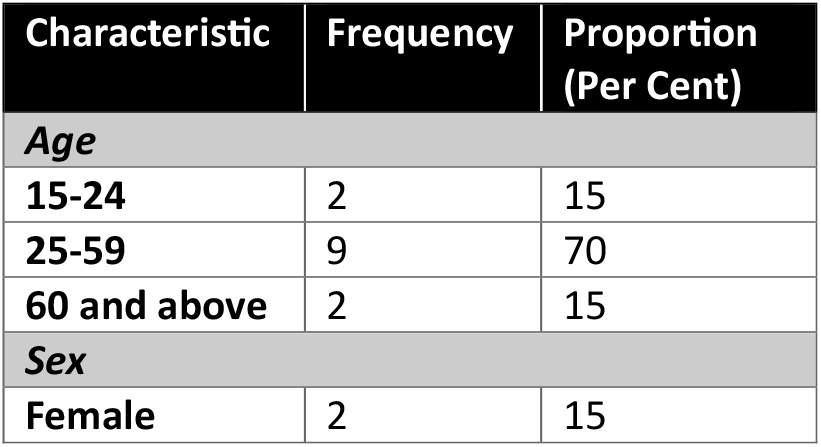
Demographic Profile of participants.

### Themes

Based on the analysis of the interview transcripts and notes four themes were identified about the PROs relevant to trauma patients in urban India. The need for full physical functioning, the need to address psychological consequences, the need to alleviate economic cost of trauma, and the need for social interactions. We have summarised the themes and sub-themes in Table 2.

**Table 2:** Themes and sub-themes.

|  |  |  |  |
| --- | --- | --- | --- |
| **1. Need for full physical functioning**<br>1.1. Reduce pain<br>1.2. Improve mobility<br>1.3. Ability to self-care and do daily activities | **2. Need to address psychological consequences**<br>2.1. Mental distress to self<br>2.2. Mental distress to those around them | **3. Need to alleviate economic costs**<br>3.1. Return to work<br>3.2. Consequences of economic burden | **4. Need for social interaction**<br>4.1. Support of family and friends<br>4.2. Need for social participation |

#### 1. Need for full physical functioning

The participants described that being able to physically function like their pre-trauma life was a major challenge they faced and was critical to their recovery.

##### 1.1. Reduce pain

Experiencing physical pain after discharge was a common problem raised by almost all the participants. Most of them felt it was excruciating at the beginning of their discharge period affecting their ability to move.

*“Some days I would be in so much pain that I would shut all the doors of the house and scream.”*

-Limb amputee participant

While it reduced over time, nearly three-fourths of them still felt some form of pain. For most of them just the pain was focused around the injured body part or when they performed a certain action, but for others it was still continuous, and at times debilitating pain.

*“(The) pain is there, but only when I am using the washroom, when the pressure builds up, or else not much pain is left.”*

-Abdominal injury participant

*“I feel continuous severe pain on my head, all the time.”*

-Traumatic brain injury participant

Whether temporary or permanent, the participants described that the pain was one of the biggest barriers for them to recover and get on with their lives.

*“If my headache is cured properly, that will be the best way for me to get better.”*

-Polytrauma participant

##### 1.2. Improve mobility

Along with reducing pain, another important physical challenge expressed by participants was not being able to move around as well as before. This included the ability to walk as well as move the injured body part.

*“I can only walk for 10-15 minutes, that’s it! If I take a chair, I can sit for only 2-3 hours! I cannot stand up more than half an hour, the maximum I have been able to stand up is half an hour!”*

-Polytrauma participant

*“Sitting is a big issue. (All I am able to do now is) bed rest. So, the entire day I have to lie in bed, so my spinal cord starts to hurt.”*

-Polytrauma participant with spinal cord injury

Though some of them have regained the ability to move around and perform physical actions as before, for others this has not happened for months after being discharged. Restricting their movement unlike before the trauma.

*“This has happened to me so many months ago and I am not able to lift my hand till now.”*

-Polytrauma participant

Some were and are still using devices like walkers and prosthetics to move. Yet, it still was challenging to move independently as before.

*“I had the most difficulty with learning to walk again on the support of just one leg. I do not wish to be dependent on a prosthetic leg or a walker, it still is so painful and difficult to walk.”*

-Limb amputee participant

##### 1.3. Ability to self-care and do daily activities

Together the pain and poor mobility impeded the ability of the participants to perform self-care activities such as bathing or using the washroom and to resume their daily chores or activities like meeting others and attending social gatherings. Many of the participants felt that it was crucial to be able to self-care and do daily activities for them to feel like they have physically recovered from the trauma.

*“I was unable to access it (toilet) due to this injury. There was a bucket, and we do not have a sitting toilet (western-style commode) so we used to throw it.”*

-Polytrauma participant

*“I needed to go to the market, I did not have anything at home, as I was the one who used to go to the market I had to call out for groceries to the kids around the neighbourhood.”*

-Polytrauma participant

*“This injury hampered my life for 2 to 3 months! I couldn’t stand for long at a stretch so I couldn’t go anywhere or do what I normally do. I have missed meeting people or being there for social gatherings.”*

-Polytrauma participant

#### 2. Need to address psychological consequences

Another key outcome that came through the interviews was psychological needs. Though the patients did not use the term “mental” or “psychological”, almost half of them described having symptoms that appeared to need psychological care. These included, being unable to sleep, constantly feeling angry, feeling alone, worrying, and unable to share problems. These in turn, deeply affected them and those around them.

##### 2.1. Mental distress to self

Participants reported that the trauma and its aftermath left them feeling emotionally disturbed during post discharge. Some reported feeling groggy and tired or unable to sleep during the recovery.

*“He keeps on saying that my mind is not working.”*

*-*Polytrauma participant

*“I cannot sleep at all since then. I am awake wondering what to do.”*

-Polytrauma participant

Others reported feeling angry and agitated since the trauma.

*“He gets very angry, irritated, keeps talking and shouting.”*

*-*Polytrauma participant

##### 2.2. Mental distress to those around them

Participants and their caregivers also reported how the trauma affected those around by making them tensed and worried about the challenges borne by the patients or the stress the patients felt.

*“He keeps on shouting, and that is why my mind also gets affected.”*

*-*Polytrauma participant family member

*“My father and mother have faced a lot of issues, they had to live with a lot of tension because of me.”*

-Polytrauma participant

These psychological stress symptoms were attributed to the consequences of the trauma such as pain, inability to move, changes in physical appearance, the ability to accomplish tasks or fulfil roles like they did before.

*“The biggest problem was that I kept thinking will my waist be ok, or no? Will I be able to walk or stand up, or no? These thoughts had taken up my mind.”*

*-*Polytrauma participant with spinal cord injury

*“I used to run the family and I now (all I do is) just sleep, who will manage now? It keeps me worrying all the time.”*

-Limb amputee participant

Moreover, not having adequate awareness or opportunities to deal with these psychological symptoms would exacerbate their mental health.

“*The patient usually has little to no emotional support and mental health awareness to be able to reach out to people during these times or feel comfortable enough to reach out to someone and share his mental pain, making him feel worse.”*

-Medical social worker

#### 3. Need to alleviate economic costs

Another recurring need that every participant reported was the economic fallout of trauma. Some felt that it was the most important challenge they faced and the one that was the most protracted to recover from.

*“The patients are normally not aware of the extent of the financial problems until much later and that then becomes the primary concern of the persons involved.”*

-Medical social worker

*“My financial situation more than my physical strength affects my life the most.”*

-Abdominal injury participant

##### 3.1. Returning to work

By far the most common economic outcome, that was pertinent following the trauma, reported by every participant was loss of employment or reduced employment opportunities. It appeared to hamper their recovery process, affected their family life, and put immense psychological strain on them.

*“I cannot earn now. I used to work as a welder. It involves lifting heavy things. (Such) heavy work I cannot do, until I get fully recovered. That means I have no work, no money till then.”*

-Polytrauma participant

*“I have two kids, a wife and mother and father, I have to take care of them too, it is very difficult work, life is somehow going on. I am lost about what to do.”*

-Polytrauma participant with spinal cord injury

*“I am scared now. My line of work makes my stomach hurt so much. I cannot work. Now I am not able to. That’s it. I am afraid now; it keeps me awake and thinking all the time.”*

-Polytrauma participant

##### 3.2. Consequences of economic burden

Many participants stated how the loss or limited employment combined with the cost of care of trauma worsened their economic conditions. This forced them to cut back on things. Some could not complete their follow-up treatment or afford medications.

*“We are poor. We have many things to worry about all the time. Now after all this, we cannot do much. We have stopped taking certain medicines.”*

-Polytrauma participant

Some had to modify their household spending to fit the limited scope of finances due to economic costs of the trauma.

*“Finance is the main problem we have; (at that time) nobody was earning in our family. I used to earn before. We went into a lock-down on money. We had to think before eating or buying anything.”*

-Polytrauma participant

Some reported taking loans to meet the increased economic burden. This extended the recovery from trauma even longer.

*“There was only one option. We had to take out a loan and then spend it on everything. We are still repaying it and will continue that for a long time.”*

-Polytrauma participant

These economic hardships contributed to the mental distress some participants experienced after trauma.

*“I feel due to financial trouble my life has changed so much. Especially, mentally it has changed me forever!”*

-Polytrauma participant

#### 4. Need for social interaction

A major set of outcomes that many patients reported as important in their recovery after discharge was social support. Be it through caregivers or through family, neighbours, and friends, support from them was an important aspect in their return to pre-trauma life.

##### 4.1. Support of family and friends

The participants detailed the various ways their social group including friends and family played an important role after discharge. They performed a range of roles from caregiving to financial support to physical assistance in moving or self-care and fulfilling the roles of the recovering trauma patient.

*“I just wanted to say that if anyone ever gets into an accident, for them their family plays a very important role, as others can leave them, but their parents will never leave them.”*

-Polytrauma participant

*“My friends have helped me and they also gave support to my family. They were there at the hospital at every moment to take care of me. I know that They will take care of everything. Even when my parents could not come, they (friends) were there with me”*.

-Polytrauma participant

*“Patients require a sense of belonging. Understanding that they are not alone in this struggle helps them”*

-Medical social worker

The presence of a caregiver was, especially, instrumental in their recovery. The absence of such a caregiver or social support was pointed as a key barrier in attaining pre-trauma life.

*“Who will take care of me? No one is there at home, to look after me and take care. It made everything more difficult.”*

-Polytrauma participant

“*Our relatives were good so they supported us, they used to get us medicines and do the treatments, but I cannot imagine someone who is not having what relatives will do?”*

-Polytrauma participant with spinal cord injury

##### 4.2. Need for social participation

Many participants also pointed out that being able to attend social gatherings was necessary for them after trauma. These were family events, festivals, or neighbourhood events. Meeting and interacting people seemed essential for the participants in their recovery process.

*“I have missed meeting people or being there for social gatherings. I missed the festival last time, it was my favourite time of the year. I was stuck lying here (on the bed).”*

-Polytrauma participant

*“I used to feel sad that I could not meet the other women (in the neighbourhood). It was such a close bond I had and the things we do together.”*

-Polytrauma participant

## Discussion

In this paper we examined PROs considered to be most relevant for trauma patients after discharge in urban India. We wanted to understand the participants’ perspectives on which outcomes affected them the most during their recovery process. Based on their responses about their lived experience of post-discharge life and their involvement working with trauma patients, we identified four themes that we could classify the outcomes that were stated as important. Participants interviewed in this study reported the need for full physical functioning, need to address psychological consequences, need to alleviate economic cost, and need for social support as outcomes most significant to them post discharge.

According to the participants in this study, reducing pain, ability to move, and performing self-care and daily activities were important for physical functioning after trauma. Participants reported having acute pain initially that reduced over time, and some continued to experience pain long after discharge. Acute pain during the initial discharge phase that decreases over time has been documented in previous studies, including in urban India itself (37,38). There is evidence that in up to half of all trauma patients this pain can continue for months after the injury (39–41). The participants interviewed mentioned that being able to move their injured body parts and themselves as whole, like before the trauma, was a major challenge. This challenge has been demonstrated in research from other settings (42–45). The interviews also highlighted that the pain and limited mobility affected the ability of the participants to perform self-care activities like bathing or using the toilet and carry out their daily activities. The restrictions to self-care and not being able to do daily activities has also been noted in trauma literature (4,46–48). Therefore, physical functioning such as pain, mobility, self-care, and daily activities are important outcomes among post-discharge trauma patients.

Participants described having symptoms similar to what may be classified as psychological distress: insomnia, anger, worrying, and feeling tired. Studies have shown how trauma with its functional and economic consequences is linked to psychological distress symptoms like sleep disorders and tiredness as well as anxiety and depression (49–53). Apart from the post-traumatic stress disorder (PTSD), due to the event resulting in the trauma, chronic physical pain, functional limitations, economic costs of the trauma, and loss of traditional social roles are drivers of this distress (54–57). The participants also noted that the symptoms of their mental distress would often affect those around them. Studies report how those living with trauma patients themselves are affected psychologically as they are forced to bear the additional responsibility of caregiving, providing physical and financial support, and adjust to the altered way of living with a trauma patient (58–61). Thus, psychological outcomes are relevant in studying trauma care.

All the participants described how trauma posed a major economic burden in their lives. The out-of-pocket costs, both direct and indirect, especially in LMICs like India, have been well established in literature (62–65). Being able to return to some form of work was one of the critical outcomes within the economic challenges stated by most participants. Loss of employment and limited employment opportunities have been noted extensively in trauma research as outcomes held important by trauma patients during their recovery (54,66–68). Returning to work is necessary to bear the costs of trauma care, prevent impoverishment as well as is also attached to being able to participate and contribute to self and society meaningfully (69,70). Participants also spoke about how the consequences of the costs of trauma including loss of employment affected their spending and contributed to their mental stress. This echoes with research that indicating being able to return to work after trauma is associated with reduced depression symptoms (71,72). It would be useful to measure economic burden, especially return to work, as an important outcome for trauma patients.

The participants spoke about how social interaction, both in the form of social support, especially in caregiving, and social participation was imperative and valuable in their recovery after being discharged. Social support has been considered in research as an important outcome in determining recovery of trauma patients (73–75). It can be in the form of physical care, financial aid, or emotional relief (76–78). The role of caregivers, especially in settings with limited affordable and available rehabilitative services, has been recognized as being particularly vital. Being able to participate in social and community settings, is a significant step in the process of trauma returning to their pre-injury lives (54,79–81). Given that these social interactions play a role in reducing the adverse consequences of trauma, it would be of key importance in trauma research to measure them.

## Methodological Considerations

To our knowledge this paper is one of the first studies that tries to understand PROs most relevant to trauma in urban India using qualitative methods. Even though we tried to make the participant sample representative of trauma patients from different demographic and injury groups, we were not able to cover groups like older females, transgenders, burns, and paraplegia or quadriplegia. Our participants belong to two cities—Mumbai and Kolkata—and cannot fully represent other urban populations in the country. The interviews were conducted at a single point of time, indicating the participants’ experience up to that time. Following up with more interviews at different points after discharge may have captured a more comprehensive view of relevant PROs.

The data collectors, translators, and the research teams’ biases and perspectives could have influenced the interpretation and analysis process. We tried to reduce this by following the principles and best-practices recommended while conducting semi-structured interviews and performing thematic analysis. Moreover, we regularly discussed and reviewed the data collection process. Some of the interviews were conducted over the telephone, which limits the level of interactions between the interviewee and interviewer. But we wanted to respect the decision of the participant to speak over the phone and ensure their convenience. Similarly, conducting the interviews at the homes or offices of the participants, could have influenced their responses due to the presence and distractions of family members and colleagues. However, we believe that this was done to prioritise the comfort of the participants.

## Conclusion

The findings of this paper add to current knowledge of PROs relevant to discharged trauma patients in the context of urban India in particular and LMICs in general. Our study shows that the need for full physical functioning, the need to address psychological consequences, the need to alleviate economic cost, and the need for social interactions are reported to be most relevant to trauma patients in this context. This can help researchers and clinicians, working in urban India and other similar settings, to identify appropriate tools that can be used to measure PROs related to these themes for research aimed at improving trauma care and patient outcomes. Future research should explore PROs relevant for specific demographic and injury groups in different settings in India and other LMICs.

## Data Availability

All data produced in the present study are available upon reasonable request to the authors

## Acknowledgments

The authors would like to express their deepest gratitude to Anna Aaroke and Tamal Khan, from Doctors For You for being part of the data collection process for this study.

## Funding

The study was part of the Trauma life support training Effectiveness Research Network (TERN) Project funded by Doctors For You, India through grants awarded to Karolinska Institutet, Sweden by the Swedish Research Council (grant number 2020-03779) and the Laerdal Foundation (grant number 2021-0048). The funders had no say in the planning or outputs of this study.

## Ethics approval and consent to participate

Ethical approval obtained for the study from the five health facilities in this study: (Approval No: HIEC/003/FEB-22/0/RP/03/02-22, Dated: 11 Feb 2022; IEC(II)/OUT/134/2022, Dated: 8 Feb 2022; Ref No.MC/KOL/IEC/NON-SPON/1217/11/21, Dated: 31 Nov 2021; NRSMC/IEC/93/2021, Dated: 29 Dec 2021; IEC/214/22 (Ref: IEC/40/22), Dated: 20 May 2022)). Informed consent was solicited and recorded from all the participants before being interviewed.

## Disclosure statement

No potential conflict of interest was reported by the author(s).

## Notes

### Competing Interest Statement

The authors have declared no competing interest.

### Author Declarations

Institutional Ethics Committee of H.B.T. Medical College & Dr. R.N. Cooper Municipal General Hospital, Mumbai; Institutional Ethics Committee-Relating to Biomedical and Health Research (BHR), Seth GS Medical College and KEM Hospital, Mumbai; Institutional Ethics Committee, Medical College Kolkata, Kolkata; Institutional Ethics Committee, Nilratan Sircar Medical College & Hospital, Kolkata; Institutional Ethics Committee-Human Research, Lokmanya Tilak Municipal Medical College & General Hospital gave ethical approvals for this study.

### Summary of Updates

Figure 1 revised to Table 2; An author added

