## Supplementary Materials for "Patient-reported outcomes relevant to post-discharge trauma patients in urban India"

a. **Interview Guide**

1. **Patient**

| Topic | Questions/Information to be collected |
|---|---|
| Background | Age, gender, socioeconomic details (education, work), family (married, widowed, living with parents/children/spouses/alone), household conditions (type of house: slum, building, attached/communal toilet)<br><br>Could you describe what happened to you?<br>(Mechanism of injury, body part injured)<br><br>How long were you in the hospital?<br>(Duration, discharge date) |
| Challenges | Could you tell us a bit about your experience at the hospital?<br><br>What do you feel the hospital staff did or can do to make your hospital stay better?<br><br>Could you describe the physical problems you faced after the injury?<br>(Mobility, pain,<br>self-care: eating, bathing, changing, excretion, etc.,<br>daily activities: travelling, shopping, going outside the house, etc.)<br><br>Do you have any of these problems related to the injury now? Could you describe them?<br><br>Of the problems you have mentioned What do you feel was the biggest physical problem you faced?<br><br>Could you describe how the injury affected your social life?<br>(Relationship of family and friends, attending community or religious functions)<br><br>What do you feel was the biggest problem that affected your social life?<br><br>What is the nature of help you received from your family, friends, and society after the injury?<br>(caregiving, help in self-care and daily activities, or financial)<br><br>Of the types of help you have mentioned, what do you feel was the help that you needed the most?<br><br>How has your injury affected your work or income?<br>(Paid and unpaid: household chores; time to get back, difficulties in performing work)<br><br>What, if any, changes have there been in the employment of other members of your household because of the injury? |

| | |
|---|---|
| | (Quitting job or taking up a new or extra job)<br><br>What are other problems relating to the injury which affect your life currently?<br><br>How do you feel your life has changed since the injury? How has the injury affected you overall? |
| Priority areas | Of all the problems and challenges you described, because of the injury, which are the ones that you feel were the biggest?<br><br>Which of these problems do you feel affected your life the most?<br><br>Which of these problems would you say had/has to be reduced or improved for you to return back to normal? |

2. **Participant working with injured patients**

| Topic | Questions/Information to be collected |
|---|---|
| Background | Age, gender, organization, designation<br><br>How long have you worked or been involved with injured patients? (Duration)<br><br>Could you describe the nature of your work with injured patients? |
| Challenges | Could you describe a bit about the main problems injured patients face?<br><br>(Physical: Mobility, pain, self-care: eating, bathing, changing, excretion, etc., daily activities: travelling, shopping, going outside the house, etc.)<br><br>Social: Relationship of family and friends, attending community or religious functions, support<br><br>Economic: effect of injury on employment, out-of-pocket expenditure<br><br>Others (emotional, etc):<br><br>Could you share a few instances of your experience with injured patients dealing with the problems you have just mentioned? (At least 2 examples)<br>(Background of patient, type of injury, specific problems faced) |
| Priority areas | Of all the problems and challenges you described, which injured patients face which are the ones that you feel affects their life the most?<br><br>Which of these problems would you say had/has to be reduced or improved for them to return back to normal? |

b. **Themes, sub-themes, and categories**

| Themes | Sub-themes | Categories | Number of Codes |
|---|---|---|---|
| Need for full physical functioning | Reduce pain | Excruciating pain | 6 |
| | | Pain was a big challenge | 15 |
| | | Continuous pain | 3 |
| | Improve mobility | Inability to walk | 8 |

| | | | |
|---|---|---|---|
| | | Inability to sit or sleep | 5 |
| | | Depending on prosthetics to walk | 3 |
| | Ability to self-care and do daily activities | Inability to use the toilet | 5 |
| | | Inability to use the bathe | 4 |
| | | Inability to do daily activities | 6 |
| | | Inability to go college | 2 |
| | | Inability to shop | 3 |
| Need to address psychological consequences | Mental distress to self | Worry about recovery | 6 |
| | | Feeling lost | 3 |
| | | Anger issues | 4 |
| | | Insomnia | 3 |
| | | Worry about loss of role as provider | 3 |
| | | Mental fog | 2 |
| | | Need to talk to someone | 4 |
| | Mental distress to those around them | Affected by anger of patient | 3 |
| | | Worry about loss of patient's role | 2 |
| | | Worry about patient | 4 |
| | | Sad about adjusting | 1 |
| Need to alleviate economic costs | Return to work | Loss of employment | 12 |
| | | Inability to work | 8 |
| | | No choices to work | 3 |
| | Consequences of economic burden | Cannot afford treatment | 6 |
| | | Long term burden | 4 |
| | | Money needed for recovery | 2 |
| | | Mental distress due to costs | 5 |
| Need for social interaction | Support of family and friends | Need for caregiver | 15 |
| | | Family support | 14 |
| | | Help from friends | 8 |
| | | Help from neighbours | 8 |
| | | Need for family acceptance | 4 |
| | Need for social participation | Missing social functions | 6 |
| | | Missing festivals | 3 |
| | | Missing company | 8 |
